# Supervised land- and water-based exercise intervention in women with fibromyalgia: CERT- based exercise study protocol of the al-Ándalus physical activity randomised controlled trial

**DOI:** 10.1101/2024.01.24.24301515

**Authors:** Inmaculada C. Álvarez-Gallardo, Blanca Gavilán-Carrera, Ana Carbonell-Baeza, Víctor Segura-Jiménez, Daniel Camiletti-Moirón, Milkana Borges-Cosic, Virginia Aparicio, Manuel Delgado-Fernández

## Abstract

**Background:** Exercise is recommended for managing fibromyalgia; however, the scant details provided about exercise programs (EP) in the available literature make standardization, replicability, and interpretation of results difficult. The aim of the present report is to provide a comprehensive CERT (Consensus on Exercise Reporting Template)-based description of the rationale and details of the land- and water-based EP implemented in the al-Ándalus Randomized Controlled Trial (RCT).

**Methods:** Women aged 35-65 with fibromyalgia (n=180) were planned to be recruited in Southern Spain (Andalucía). The study design was composed of three groups: the usual care (control) group, the land- and the water-based supervised exercise intervention groups (n=60 for each group). Participants allocated in the exercise intervention groups undertook a 24-week supervised multicomponent (strength, aerobic and flexibility) EP (three non-consecutive sessions per week, 45-60 min/session). The rationale of the exercise program is described in detail following the CERT criteria detailing its 16 key items.

**Discussion:** This study details the supervised EP of the al-Ándalus RCT project, which may serve: 1) exercise professionals who would like to implement an evidence-based supervised EP for people with fibromyalgia in land- and water-based settings, and 2) as an example of the application of the CERT criteria.

**Trial registration:** ClinicalTrials.gov ID: NCT01490281

## INTRODUCTION

Fibromyalgia syndrome is a common[1] and complex chronic pain condition[2]. Individuals with fibromyalgia manifest musculoskeletal pain, fatigue, nonrestorative sleep, and physical and psychological impairment, among other symptoms[2, 3]. It has a substantial impact on the quality of life (QoL)[4] and results in a 5-fold higher healthcare expenditure[5]. Its diagnosis and management are often challenging because of the heterogeneous manifestations and differences between individuals with respect to dominant symptoms and their severity[6]. Exercise has been demonstrated to be a safe and effective non-pharmacological approach for managing fibromyalgia symptoms[7, 8], with the greatest impact on improving pain and symptom severity.

Multicomponent exercise interventions (i.e. a combination of aerobic, resistance and flexibility) might offer unique advantages beyond those derived from interventions employing only one type of exercise[9]. Multicomponent exercise interventions in fibromyalgia have been typically carried out in land- and water-based settings. Of noting, the physical and mechanical characteristics of each setting may elicit different physiological and biomechanical responses. Previous evidence comparing the effects of exercise in both settings concluded that similar results for overall well-being, physical function, pain, stiffness[10], and fatigue[11] were obtained under both conditions, and only a moderate difference was detected in strength, favoring land-based training[10]. However, the evidence available is limited and rated as low-quality[12], which prevents establishing firm conclusions and advising patients on a specific setting for exercise. Therefore, a better understanding on the comparative effectiveness of land- and water-based exercise interventions in fibromyalgia is relevant from a research and clinical perspective. Since a water-based approach is by far more expensive and many communities do not have this facility, comparing the benefits from different exercise settings is also relevant from a financial and social point of view.

Importantly, exercise-based treatments have shown small to moderate effects with considerable variation in the way interventions are designed and delivered[9]. In addition, evidence is usually based on low-quality studies[13] and low therapeutic validity of exercise intervention programs[12], mainly due to incomplete descriptions of exercise interventions and adherence[12]. The absence of explicit descriptions of the methods and components of the exercise interventions hinders replication, limiting our ability to assess the effectiveness of different programs[14]. The Consensus on Exercise Reporting Template (CERT) was created to standardize the reporting of exercise intervention programs[15]. The Consensus, developed by a meta-epidemiological review of exercise interventions for chronic health conditions[14], guides on several key items required to report replicable exercise programs. To translate research findings into meaningful clinical interventions and define future disease-specific recommendations, properly designed and high-quality intervention studies are needed.

The al-Ándalus study is a randomised controlled trial (RCT) to investigate the effects of 24-week land- and water-based supervised exercise interventions on disease impact, tenderness, body composition, functional capacity, QoL and cognitive function in female patients with fibromyalgia. In view of the need to explain in detail the physical exercise intervention programs for people with fibromyalgia, the present report aimed to provide a comprehensive CERT-based description of the rationale and details of the 24-week land- and water-based exercise program implemented, which complements the previously published study design of the al-Ándalus RCT[16].

## METHODS /DESIGN

### Study design

The study design of the al-Ándalus physical activity RCT (ClinicalTrials.gov ID: NCT01490281, 12/12/2011) has been published previously elsewhere[16] and as part of a doctoral thesis[17]. Briefly, the required sample size was determined for the primary outcome variable, i.e. overall score of Fibromyalgia Impact Questionnaire (FIQ)[18]. As a result, a minimum of 60 women with fibromyalgia were needed for each group. As the study design was composed of two types of interventions (land-based and water-based), we planned to recruit a total of 180 women with fibromyalgia (i.e., two supervised intervention groups and one usual care control group of 60 participants each). The Medical Ethics Committee of Hospital Virgen de las Nieves (Granada, Spain) approved the study design, study protocols and informed consent procedure[17].

### Consensus on Exercise Reporting Template (CERT)

The CERT ‘Explanation and Elaboration Statement’[15] was used to ensure the methodological quality of the al-Ándalus trial. The CERT is a 16-item checklist designed to evaluate the completeness of exercise intervention reporting and spans ‘who’, ‘what’, ‘when’, ‘where’, and ‘how’. The rationale of the exercise program implemented in the al-Ándalus trial will be described following the CERT criteria recommendations for detailing the 16 items (Table 1).

**Table 1.**
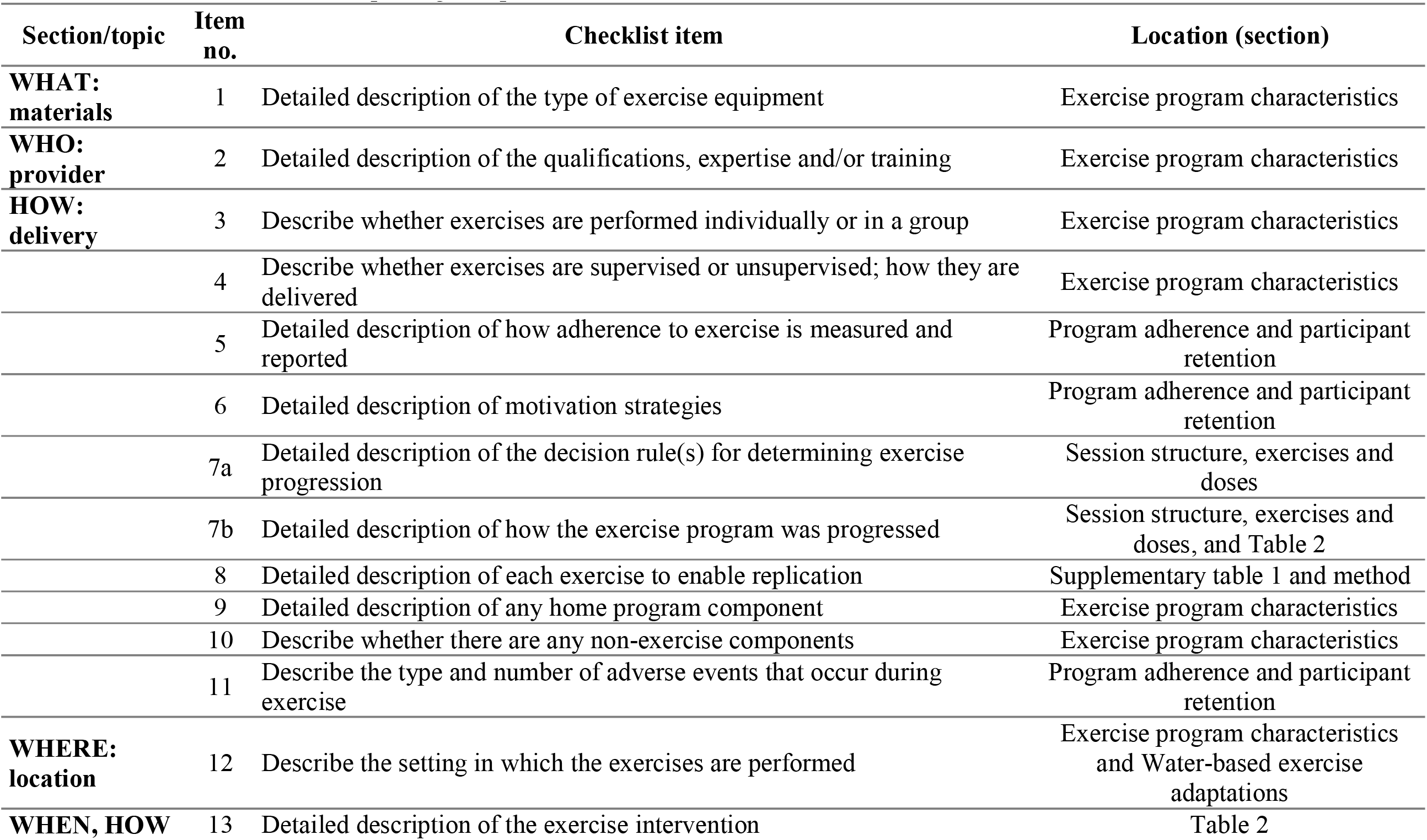
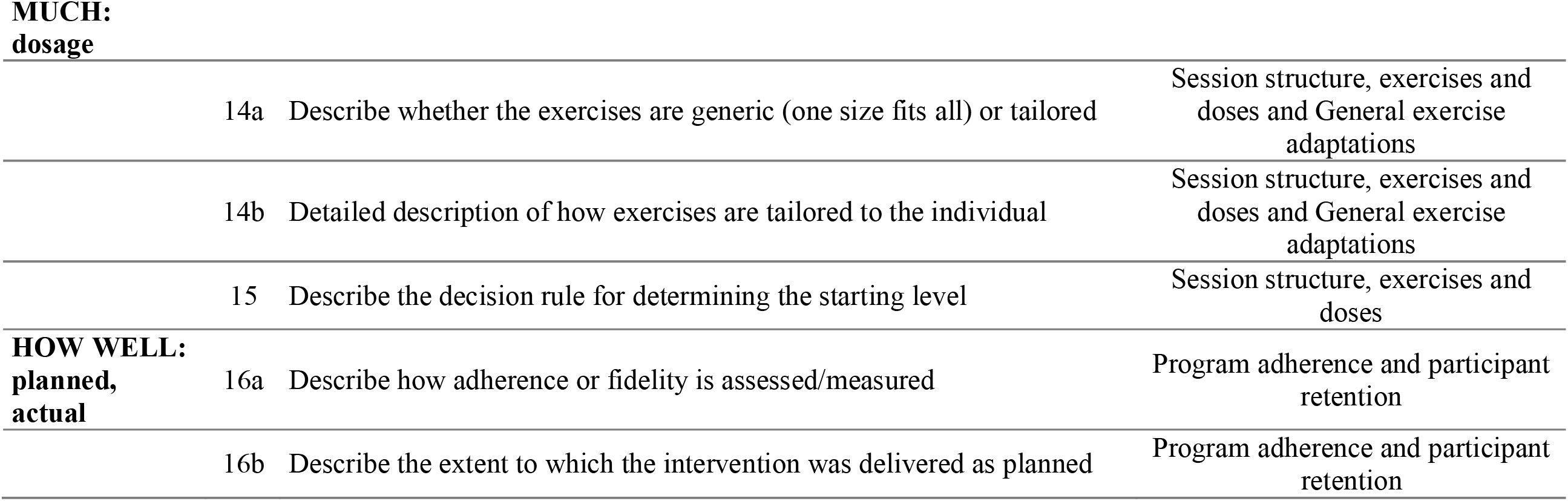
Consensus on Exercise Reporting Template (CERT) checklist from the al-Ándalus exercise intervention trial.

### Program structure

#### Exercise program rationale

The land- and water-based supervised exercise intervention groups were trained on three non-consecutive days/week (45-60 min per session) over a 24-week period (72 sessions in total) following the same exercise protocol in both groups. A six-month program was planned because previous studies[19, 20] evidenced that more than 12 weeks of exercise were needed to observe cumulative changes in pain and other fibromyalgia symptoms. Following the American College of Sports Medicine (ACSM) recommendations[21], the exercise program was performed 3 days per week on alternate days (Mondays, Wednesdays and Fridays) to provide the necessary between-session resting time.

The intervention aimed to improve cardiorespiratory fitness, muscle strength, and joint range of motion, which are inversely associated with disease severity[22] and fibromyalgia symptomatology[23–29]. For this purpose, we used a supervised multicomponent exercise program (i.e., an exercise program combining aerobic, resistance and flexibility training). The initially planned exercise interventions met the minimum training standards of the ACSM for patients with fibromyalgia[21]. During their involvement in the study, all participants continued to receive standard care, primarly consisting of pharmacological treatment, from their healthcare providers.

#### Exercise program characteristics

The interventions were supervised and performed in 17 waves from 2011 to 2013, between November and May. All training sessions were conducted in fitness centers or patient association facilities (Item 12). The exercise equipment needed (Item 1) included chairs, fitness mats, tennis balls, elastic bands (low and medium resistance), and light dumbbells for the land-based exercise program, and flotation materials such as pool noodles, pulls, float boards, and tennis balls for the water-based exercise program. Music and stereo equipment were used in all sessions. The program was supervised (Item 4) and there were no other home programs (Item 9) or non-exercise components (Item 10). The patients were organized into groups of 7-15 women (Item 3), each supervised by the same instructor throughout the exercise program. The instructors were certified Sport Science professionals (Item 2) who performed program-specific training before the intervention. Moreover, to standardize the implementation of the intervention program, an operating manual was developed with detailed guidelines. A total of 13 instructors monitored 17 different exercise groups across 9 cities in Andalusia, located in Southern Spain.

A full description of the training periodization (Item 13) with a detailed summary of the general principles for exercise prescription (type, duration, frequency, intensity, volume, and mode of exercise) for each session is presented in Table 2[17].

**Table 2.**
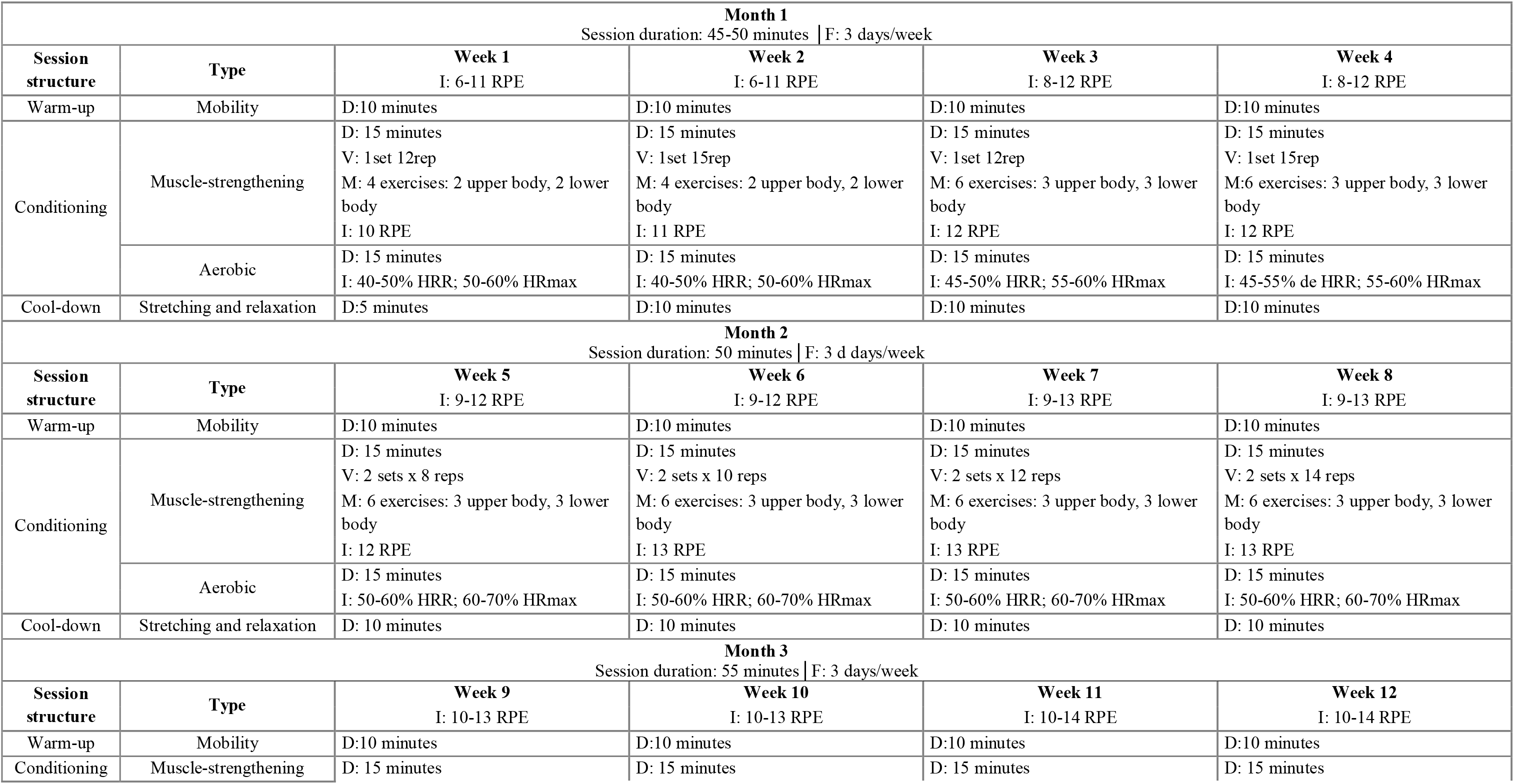
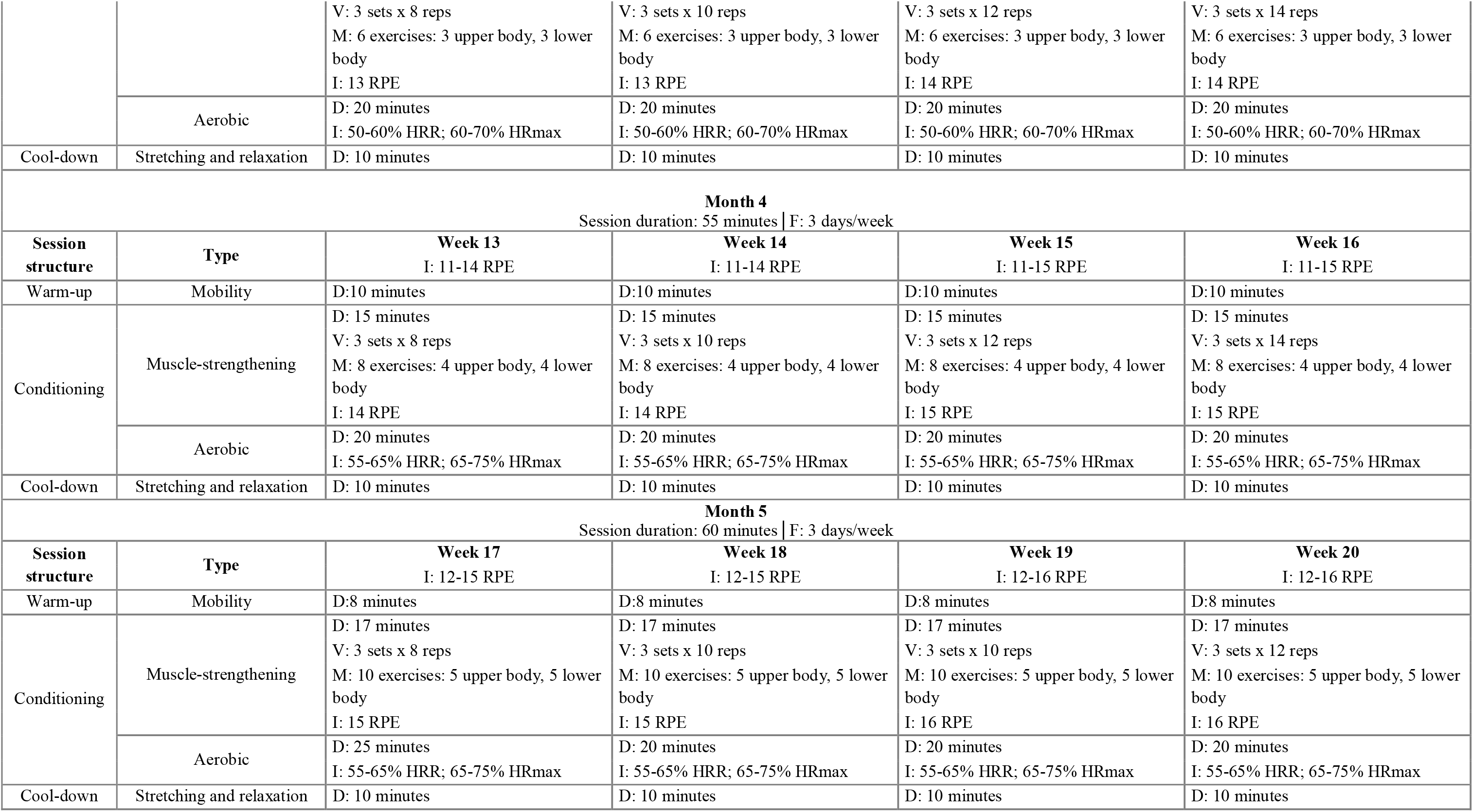
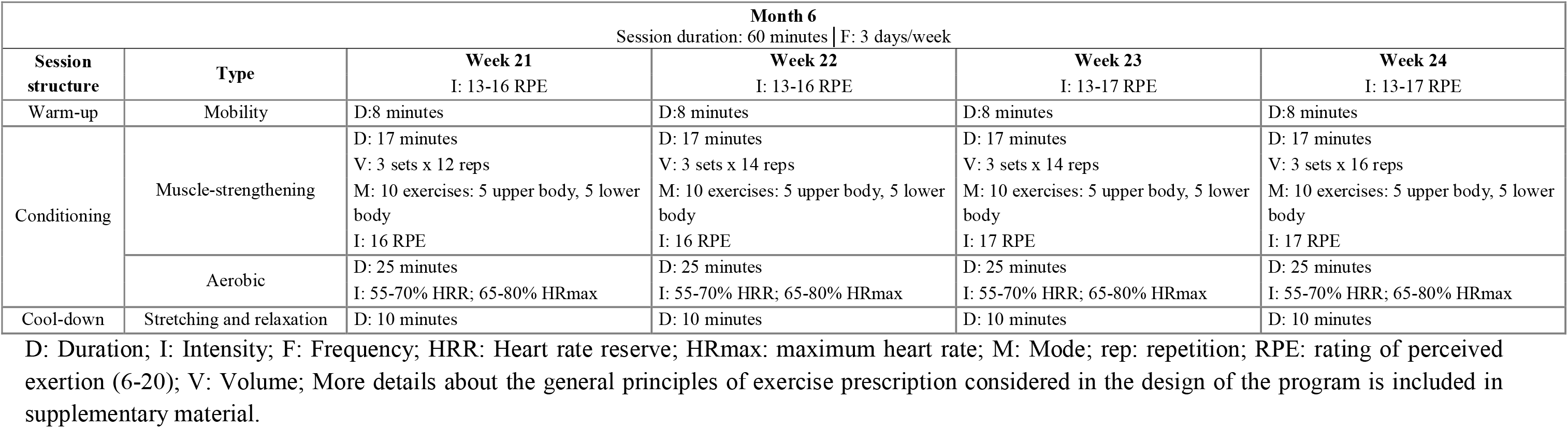
Summary of the exercise progression planned for the al-Ándalus exercise intervention trial.

#### Session structure, exercises and doses

Each session lasted from 45 min (week 1) to 60 min (week 24) and was divided into the following parts: warm-up (8-10 min), conditioning [muscle-strengthening (15-17 min) and aerobic (15-25 min)], and cool-down (10 min). During the first few weeks, the main part of the sessions focused on exercise familiarization and learning of basic movement patterns. The following is a more detailed explanation of the work carried out in each part of the session:

##### Warm-up

Each session included an 8-10 minutes warm-up with joint mobility exercises and slow walking with global movements. Mobility exercises included mobilisation of the main joints of the body (neck, shoulders, elbows, wrists, hands, hips, knees and ankles) of 6-8 repetitions each at a slow speed. They were performed starting with the upper body (neck) and progressing to the lower body. The same sequence of movements was repeated in all sessions to help the participants become familiar with them. After joint mobility, slow walking with global movements was included to increase body temperature and blood flow. Different types of coordinated displacements with arm movements were used. These warm-ups were accompanied by the use of music.

##### Conditioning

The main part of the session consisted of resistance strength circuit training followed by aerobic training. Resistance strength circuit training comprised a whole-body exercise routine involving major upper and lower body muscle groups to improve performance in functional activities. A detailed description of the exercises used in the al-Ándalus physical activity randomized controlled trial (Item 8) is provided in Supplementary Table 1. Strengthening exercises included both single- and multi-joint exercises such as adapted push-ups, bench presses, pulls, biceps curls, shoulder exercises, hip mobility, glute kickbacks, stand-ups from the seated position, semi-squats, deadlifts, lunges, leg extensions, and standing calf raised (similar exercises with slight variations). The Borg Rating of Perceived Exertion (RPE) scale was used to monitor intensity. The planned intensity ranged from 10 RPE (week 1) to 17 RPE (week 24) (RPE 6-20 scale). All major muscle groups were exercised using movements that involved minimal work above the head, and were performed near the midline of the body. The starting level and progression were determined following the ACSM recommendations for exercise training for people with fibromyalgia[21] and our previous experience with this population[30–32] (Items 6 and 7a). The speed of concentric contractions was low, and eccentric and isometric contractions were maintained at a minimum. The starting level (week 1) included 1 set of 10-12 repetitions covering 4 exercises and progressively increased each week until the final level (week 24), which included 3 sets of 16 repetitions covering 10 exercises (see Table 2). Progression was achieved by first increasing the volume (progressively increasing the number of exercises, sets, and repetitions) and then the intensity (increasing load/resistance, movement speed, and reducing recovery time) (Item 7b). The exercises progressed in intensity based on the participant’s response to them and daily fibromyalgia symptoms to reach the RPE set for each session (Items 7b, 14a and 14b). Lower- and upper-limb exercises were alternated, and periods of active resting were included between each set adjusted to the participantś perceptions. The participants were instructed to exercise through the full range of motion and to avoid the Valsalva maneuver.

Resistance strength training was followed by 15-25 minutes of aerobic training, which was planned to progress from 40-50% Heart Rate Reserve/50-60% Maximum Heart Rate (at the beginning of the intervention) to 55-70% Heart Rate Reserve/65-80% Maximum Heart Rate (in the last month of the intervention). Low-impact exercises involving large muscle groups were performed (e.g., aerobic circuits, dancing, and games involving displacement and walking at different speeds). As before, progression was achieved by first increasing the volume (time) and then the intensity (Item 7b)[17].

Finally, each session ended with a 10-minutes cooling down period with static stretching, holding the stretch for 10-30s (to the point of gentle tension), and relaxation exercises. The stretches involved the neck, shoulders, chest, arms, lower back, upper back, hands, glutes, hamstrings, and calves. Stretches were individualized to teach each patient how to avoid overstretching. Progressive relaxation included guided imagery with breathing awareness, diaphragmatic respiration, progressive muscular relaxation, and adapted contraction–relaxation techniques[17].

#### General exercise adaptations

Exercises were modified or adapted if the participants experienced exacerbation of symptoms. The program was tailored to the individual, depending on the severity of the fibromyalgia, using perceived exertion (RPE) (Item 14a). The Borg’s conventional scale (6-20 points)[33] was used. Participants were informed of the target RPE for each session and during the session, they were motivated to reach it, particularly in terms of resistance strength and aerobic training (Item 14b). Initially, the exercise was carried out at a physical exertion level that the participants were able to do without undue pain or exacerbation of other symptoms and slow progression to allow physiological adaptation without an increase in symptoms. Pain pre- and post session (using a 0-10 cm VAS scale), as well as the RPE post-session, were assessed using a diary to determine the ongoing overall impact of the disease and symptoms of fibromyalgia with exercise.

To determine adherence to the intensities of the planned exercise program, each participant indicated an overall RPE after each session through a diary. In addition, the heart rate was controlled using a monitor (Polar RCX-3, Kempele, Finland) worn by a subsample in each group of three people, alternating the participants every three sessions (on fridays, progressions were established weekly) (Item 5).

#### Land-based exercise adaptations

In the first weeks, some exercises were performed in a seated position to avoid fatigue, and participants were encouraged to perform the exercises standing when possible. To increase the load and achieve the set RPE, elastic bands and dumbbells (0.5-2 kg) were added progressively.

#### Water-based exercise adaptations

The pool had to meet the following characteristics for the water-based exercise intervention groups: depth of ∼120 cm (chest-high) and water temperature of ∼30°C (Item 12). Water properties were considered for the water-based exercise intervention program. Movements were performed at a slow pace, aquatic sports equipment was used to increase the resistance or as an aid, stretching was performed in the standing position, and relaxation was performed while floating. The intensity of the training and the muscle groups activated were kept as similar as possible to those of the land-based exercise intervention program.

#### Usual care group (the control)

Participants assigned to the usual care (control) group were placed in the waiting-list regime. They received general information about the disease and were given leaflets containing information about the benefits of being physically active and general guidelines about how to increase their daily physical activity levels. After the follow-up assessment, participants were allowed to perform the exercise program (Item 16a).

#### Program adherence and participant retention

Attendance and reasons for not attending each session and dropouts were recorded by the instructors. To maximize participant adherence and retention, several strategies were implemented: group sessions were performed[21], music was used during the sessions, participants’ preferences were taken into account when possible (for example, by including dance activities in the aerobic part of the sessions), individualized attention was given with exercises tailored to participants by instructors, and phone calls were made after missed sessions (item 6). Phone calls also allowed us to identify and monitor non-attendance due to adverse events. Training adherence was defined as the number of sessions attended divided by the number of prescribed sessions (n=72). Achieving ≥70% adherence at all planned training sessions was considered a successful completion and adherence rate.

## DISCUSSION

The present work describes the exercise program of the al-Ándalus RCT in detail using the CERT tool. The al-Ándalus RCT study aims to investigate the effects of 24-week land- and water-based exercise interventions on disease impact, tenderness, body composition, functional capacity, QoL, and cognitive function in female patients with fibromyalgia. Current research indicates that physical exercise is an effective treatment for managing fibromyalgia symptoms[9, 34, 35]. However, a shortcoming often found is the lack of details regarding the programs employed, highlighting the need for standardized reporting. The inclusion of a detailed description of key exercise variables (i.e., frequency, duration, intensity, progression, and type) is strongly encouraged [7, 36] in order to: i) increase reproducibility, ii) increase the generalizability of study findings (if the intervention is well-described, other researchers can better apply the intervention to different groups, settings, and circumstances), iii) interpret the results (accurate descriptions of exercise interventions allow for a more precise interpretation of study results), iv) improve clinical practice and v) define future disease-specific recommendations [12]. All these key exercise variables have been described in the present work through the CERT template for the exercise program of the al-Ándalus RCT.

The exercise intervention program was designed to meet the minimum training standards of the ACSM[21, 37]. Previous studies implementing exercise interventions in people with fibromyalgia and our experience working with this population[31, 32, 38, 39] were also considered for the design. Traditionally, aerobic or muscle strength training have been the most investigated exercise modalities in fibromyalgia[7]. However, multicomponent exercise interventions might offer unique advantages beyond those derived from interventions employing only one type of exercise[9]; therefore, this type of training was implemented in the al-Ándalus trial. It is worthe noting that the same multicomponent exercise program was designed for land- and water-based intervention groups, with an adapted version to the restrictions and peculiarities imposed by water. Therefore, the exercise intervention program designed here maximized: i) impact on different physical fitness components, and ii) comparability between settings.

Whilst there is good evidence of the short-term benefit of exercise, long-term interventions studies are scarce. In a previous meta-analysis[9] about the effects of multicomponent exercise in fibromyalgia symptomatology, the mean duration of the analyzed exercise programs was 12 weeks (ranging 6 to 26 weeks). Importantly, in a previous study carried out by our research group[19], it was found that exercise interventions of more than 12 weeks are needed to obtain cumulative changes in pain[19]. Thus, we decided to plan a 24-week exercise program. In addition, long-term benefits in fibromyalgia after exercise cessation are unclear due to the relative lack of follow-up, limited length of follow-up, or limited follow-up phase information of the available studies[9]. Therefore, the inclusion of a 12-week follow-up in the al-Ándalus trial will help to better understand the persistence of the changes provided by multicomponent exercise in different settings.

When the exercise program for the al-Ándalus RCT study was designed, the intention was to create an affordable, low-cost program maximizing its replicability and accessibility. Thanks to these efforts, it has been possible to carry it out in 17 different exercise groups located in 9 cities. However, water-based exercise programs are more expensive and less accessible than land-based programs due to the need for swimming pool facilities. In addition, most community swimming pools are heated between 26°C and 28°C. However, the more suitable swimming pools for people with fibromyalgia[10] are those pools for therapeutic purposes, which are usually heated to between 30°C and 32°C, being more expensive and less common. For this reason, it is not only of clinical and public health, but also of economic relevance to better understand whether the benefits of land-based exercise for fibromyalgia are similar to those of exercise undertaken in water.

Adherence rates are fundamental to understanding the clinical efficacy of exercise-based interventions[40] and an important factor mediating the effectiveness of an exercise program. However, there is a lack of information about adherence and monitoring in previous RCTs studies that applied exercise to people with fibromyalgia[41, 42]. The assessment and subsequent report of exercise adherence rates should be an integral component of all RCTs and review articles studying the effects of exercise interventions[34]. At the same time, adherence to exercise in fibromyalgia is complex, as some participants could perform exercise while experiencing high levels of pain while others would avoid movement because of fear of exacerbation[43]. As adherence is an agreed behaviour[44], the sessions of the al-Ándalus exercise program were tailored to the participants’ needs as much as possible, accounting for their perspectives and preferences. For example, participants were asked about their satisfaction filling a diary at the end of each session. Moreover, an exercise intervention in groups was planned because it has shown to provide a social support system for these patients, reducing physical and emotional stress and promoting exercise adherence[21]. In the al-Ándalus trial, attendance was controlled, which will allow for reporting adherence in future studies analyzing the effects of this intervention program. Similarly, thanks to the recording of the reasons for non-attendance, we can detect possible adverse effects and explore possibilities to increase adherence.

Altogether, this report presents an evidence-based supervised exercise intervention, following the CERT reporting guidelines, and a comprehensive rationale underlying each step conducted during its design. Consequently, this evidence-based report can be used for exercise prescription in women with fibromyalgia, contributing to transparent reporting of exercise interventions in people with fibromyalgia. This ensures understanding by different health professionals from multidisciplinary teams, although the exercise professionals (i.e., certified Sport Science professionals) should play a relevant role in its implementation[45].

The present study details the supervised exercise program of the al-Ándalus RCT project, which may serve: 1) exercise professionals who would like to implement an evidence-based exercise program for people with fibromyalgia in land- and water-based, and 2) as an example of the application of the CERT criteria.

The results of the al-Ándalus RCT study will be disseminated as far as possible. The dissemination strategy will use different channels to reach a large number of stakeholder groups and individuals, at the local, national and international levels; this will include dissemination in academic media, social media and print media (among others). As stated above, the al-Ándalus RCT study was designed to maximise its replicability and accessibility trying to contribute as much as possible to scientific knowledge about the effects of exercise in people with fibromyalgia and helping to improve the QoL of people that suffer it.

## DECLARATIONS

### Ethical approval and consent to participate

This study will be performed following the ethical guidelines of the Declaration of Helsinki, last modified in 2000. Ethics approval was obtained by the Ethics Committee of the Virgen de las Nieves Hospital (Granada, Spain). All participants were over 18 years old and gave written informed consent.

### Consent for publication

Persons who appear in the photos in the supplementary material gave their informed consent for publication.

### Availability of data and materials

Data and material used and/or analyzed during the current study will be made available from the corresponding author on reasonable request after the conclusion of the study.

The study results will be published both as publications and within the frame-work of national and international congresses.

### Conflict of interest

None declared

### Funding

This project was funded by the Spanish Ministry of Science and Innovation [I+D+i DEP2010-15639]. ICAG was supported by the Spanish Ministry of Economy and Competitiveness [BES-2011-047133]. BGC was supported by the Spanish Ministry of Education [FPU15/00002] and the Spanish Ministry of Universities Next Generation “Margarita Salas” Grant Program. VSJ was supported by the Spanish Ministry of Education [AP-2010-0963]. MBC was supported by the Spanish Ministry of Education [FPU14/02518]. This work was supported by a grant/ voucher (#Q322PV44) from the European Alliance of Associations for Rheumatology (EULAR). The content is solely the responsibility of the authors and does not necessarily represent the official views of EULAR

### Authors’ contributions

MDF (principal investigator), ACB and VA conceived the study. ICAG designed the exercise intervention protocol under the supervision of MDF, ACB, VA and DCM. ICAG and BGC wrote the initial draft of the manuscript. All authors critically revised and approved the submitted version of the manuscript.

## Data Availability

All data produced in the present study are available upon reasonable request to the authors

## Acknowledgements

The authors acknowledge the help of the participants that are taking part in the study.

## Abbreviations

QoL: quality of life
CERT: Consensus on Exercise Reporting Template
RCT: Randomised controlled trial
FIQ: Fibromyalgia Impact Questionnaire
ACSM: American College of Sports Medicine
RPE: Borg Rating of Perceived Exertion
VAS: Visual Analogue Scale

## Notes

### Competing Interest Statement

The authors have declared no competing interest.

### Author Declarations

The Medical Ethics Committee of Hospital Virgen de las Nieves (Granada, Spain) gave ethical approval for this work

### Summary of Updates

Extra information added in the funding section: This work was supported by a grant/ voucher (#Q322PV44) from the European Alliance of Associations for Rheumatology (EULAR). The content is solely the responsibility of the authors and does not necessarily represent the official views of EULAR

