## Supplementary table 1 for "Supervised land- and water-based exercise intervention in women with fibromyalgia: CERT- based exercise study protocol of the al-Ándalus physical activity randomised controlled trial"

| Supplementary table 1. Detailed description of the al-Ándalus muscle strengthening exercises | |
| --- | --- |
| AL-ANDALUS MUSCLE-STRENGTHENING EXERCISES (CIRCUITS) | |
| EXERCISE DESCRIPTION |  |
| UPPER LIMB | |
| **U1. Adapted push-up**  From a standing position, with the palms of the hands resting on the pool curb or a wall, perform flexion and extension of the elbows keeping the body straight.  ADAPTATIONS, VARIATIONS AND PROGRESSIONS:   a. Execution speed (from slowest to fastest) b. Positions and equipment:  - Water-based: distance of the feet from the edge of the pool (from closer to further away, i.e. inclination progression) - standing pushing water with open hands (progression to mittens) - standing pushing pool noodles - idem pushing float boards forward and to the chest. - Land-based: distance of feet to the wall (from closer to further away, i.e. inclination progression).   *SPECIAL CONSIDERATIONS:  - Pay special attention to the correct execution of the technique, be careful with hip movements. Back and hips should stay in neutral position throughout the exercise. It is recommended to make an approximation of the scapulae throughout the movement. | Please contact the corresponding author to request access to these images. |
| **U2. Adapted floor bench press**  Initial position: stand, seated or lying supine with legs bent (for land-based) and arms stretched forward slightly lower than shoulder height. Perform scapular retraction opening arms to the sides and returning to the initial position.  ADAPTATIONS, VARIATIONS AND PROGRESSIONS:   a. Execution speed (from slowest to fastest) b. Equipment:   Water-based: pushing water with open hands (progression to mittens) - pushing pool noodles.  Land-based: without equipment, with dumbbells or elastic band.  *SPECIAL CONSIDERATIONS:  - Pay special attention to the correct execution of the technique.  - In the pool, perform with water up to your neck and arms submerged. | Please contact the corresponding author to request access to these images. |
| **U3. Arms movements of breaststroke technique**  Initial position: stand or seated (land-based). Perform movement of arms of breaststroke technique without displacement of the body.  ADAPTATIONS, VARIATIONS AND PROGRESSIONS:   a. Execution speed (from slowest to fastest) b. Equipment:   Water-based: pushing water with open hands (progression to mittens)  Land-based: without equipment or with dumbbells  *SPECIAL CONSIDERATIONS:  - In the pool, perform with water up to your neck and arms submerged. | Please contact the corresponding author to request access to these images. |
| **U4. Arms movements of croll technique**  Following the same instructions as exercise U3 with movement of the arms of the front crawl technique. | Please contact the corresponding author to request access to these images. |
| **U5. Pull:**  **Land-based:** standing with knees slightly bent and arms stretched forward by gripping an elastic band. Perform scapular retraction in addition to a shoulder extension with elbow flexion, and back to the initial position. Keep arms close to the body.  ADAPTATIONS, VARIATIONS AND PROGRESSIONS:  a. If functional capacity is severely reduced, it can be adapted to a sitting position.  b. Resistance bands of different resistance, adjusted to get to work in the RPE established.  b. Adaptation with dumbbells.  c. Perform with one arm and then with the other or both arms simultaneously.  **Water-based:** adaptations by changing the resistance bands for equipment that generates resistance in the water: position of the hands, pool noodles and float boards.  ADAPTATIONS, VARIATIONS AND PROGRESSIONS:  a. Change the position of the legs by placing one in front with the knee slightly bent and the other behind to support the resistance when moving the arms.  b. Perform with one arm and then with the other or both arms simultaneously. | Please contact the corresponding author to request access to these images. |
| **U6. Shoulder abduction and adduction**  Stand or seated (land-based), perform shoulder abduction and adduction.  ADAPTATIONS, VARIATIONS AND PROGRESSIONS:   a. Perform with one arm and then with the other or both arms simultaneously b. Execution speed (from slowest to fastest) c. Equipment:   Water-based: pushing water with open hands (progression to mittens)  Land-based: without equipment, with dumbbells or resistance bands.  *SPECIAL CONSIDERATIONS:    - In the pool, perform with water up to your neck and arms submerged.  - Pay special attention to the correct execution of the technique. | Please contact the corresponding author to request access to these images. |
| **U7. Shoulder flexion**  Stand or seated (land-based), perform shoulder flexion.  ADAPTATIONS, VARIATIONS AND PROGRESSIONS:   a. Perform with one arm and then with the other or both arms simultaneously b. Execution speed (from slowest to fastest) c. Equipment:   Water-based: pushing water with open hands (progression to mittens)  Land-based: without equipment, with dumbbells or resistance bands.  *SPECIAL CONSIDERATIONS:  - In the pool, perform with water up to the neck and arms submerged.  - Pay special attention to the correct execution of the technique. | Please contact the corresponding author to request access to these images. |
| **U8. Triceps kick-back**  Land-based: Stand in the middle of the resistance band with the right foot. Hold one end of the resistance band in the left hand and take a step back with the left foot so you’re in a split stance with the right hand resting on the front knee, leaning your torso forwards so it’s at a 45° angle to the floor. Hold the upper arm by your side and make a 90° bend at the elbow. Keeping your upper arm static, extend the arm back to stretch out the band.  ADAPTATIONS, VARIATIONS AND PROGRESSIONS:   a. Without equipment, with dumbbell or resistance band, shorten the length of the band if it is needed to increase the resistance. b. If functional capacity is severely reduced, it can be adapted to a sitting position. c. It is advisable to do one arm at a time   Water-based: Same initial position and execution technique working with water resistance.  ADAPTATIONS, VARIATIONS AND PROGRESSIONS:   a. Equipment: pushing water with different position of hands and progression to mittens, looking for progression on resistance surface with water. b. Execution speed (from slowest to fastest).   *SPECIAL CONSIDERATIONS:  - It is advisable to do one arm at a time, so the person can focus on working the muscles in the right way.  - In the pool, perform with water up to the neck and arms submerged. | Please contact the corresponding author to request access to these images. |
| **U9. Biceps curl**  Standing with feet about hip-width apart or seated (land-based). Keep abdominal muscles engaged. Let arms relax down at the sides of your body with palms facing forward. Keeping upper arms stable and shoulders relaxed, bend at the elbow and lift the weights so that the hand approach your shoulder. Your elbows should stay tucked in close to your ribs and back to the starting position.  ADAPTATIONS, VARIATIONS AND PROGRESSIONS:  Start with one one arm at a time and progress to both at the same time.  Land-based:   a. If functional capacity is severely reduced, it can be adapted to a sitting position. b. Without equipment, with dumbbell or resistance band, shorten the length of the band if it is needed to increase the resistance.   Water-based:   a. Equipment: pushing water with different position of hands and progression to mittens, looking for progression on resistance surface with water. b. Execution speed (from slowest to fastest).   *SPECIAL CONSIDERATIONS:  - Don’t recruit the shoulders or torso to swing the weights up when doing the dumbbell curl.  - In the pool, perform with water up to the neck and arms submerged. | Please contact the corresponding author to request access to these images. |
| LOWER LIMB | |
| **L1. Hip Abduction**  Stand straight, legs slightly apart, shift weight to one leg and, keeping balance, perform hip abductions by lifting the other leg out to the side. To do repetitions stablished on one leg and then repeat with the same number on the other side. It should feel the exercise working on your hip abductor muscles.  ADAPTATIONS, VARIATIONS AND PROGRESSIONS:  As variation, if a pendulum-shaped movement is made, crossing the leg in front, the adductors will also be worked.  Land-based:   a. If functional capacity is reduced, hold on to something solid, such as a chair or wall. Move on to performing without support when possible. b. Without equipment, with ankle weights or resistance band. Place the band around the legs and stretch the band by lifting the leg out to the side. c. Perform the same exercise starting from the initial position lying on your side.   Water-based:   a. Use hand support on the edge of the pool if necessary. b. Execution speed (from slowest to fastest).   *SPECIAL CONSIDERATIONS:  - Done properly, all of the work should be done by your hip and glute muscles while the rest of your body should stay static.  - Pay special attention to the correct execution of the technique. | Please contact the corresponding author to request access to these images. |
| **L2. Hip mobility drawing circles with foot**  Same exercise as L1 drawing a circle in the air with the foot of the free leg.  ADAPTATIONS, VARIATIONS AND PROGRESSIONS:   a. Direction of movement of the leg: forwards or backwards. b. Same variations that L1 for land-based and water-based. | Please contact the corresponding author to request access to these images. |
| **L3. Standing Single-Legged Knee Drive**  Stand on one foot facing away from the anchor and find the balance, then extend your working leg behind you. Drive working knee up to hip height. Return to the start and repeat right away. Do all reps on one side before switching.  ADAPTATIONS, VARIATIONS AND PROGRESSIONS:  As variation, drive working with leg extended, without knee flexion.  Land-based:   a. If functional capacity is reduced, hold on to something solid, such as a chair or wall. Move on to performing without support when possible. b. Without equipment, with ankle weights or resistance band.   Water-based:   a. Use one-hand support on the edge of the pool if necessary. b. Execution speed (from slowest to fastest).   *SPECIAL CONSIDERATIONS:  - Done properly, all of the work should be done by your hip while the rest of your body should stay static.  - Pay special attention to the correct execution of the technique. | Please contact the corresponding author to request access to these images. |
| **L4. Leaning glute kickback**  Standing on one foot, lean forward, using a chair or on the edge of the pool (water-based) for support. Now kick one leg back, stretching out as far as you can. Go back to the starting spot.  ADAPTATIONS, VARIATIONS AND PROGRESSIONS:  Donkey kickbacks, kneeking glute kickback (land-based).  Land-based:   a. Without equipment, with ankle weight or resistance band. Place the band around the legs and stretch the band by lifting the leg out to the behind.   Water-based:   a. Execution speed (from slowest to fastest).   *SPECIAL CONSIDERATIONS:   - Done properly, all of the work should be done by glute muscles while the rest of your body should stay static.   - Pay special attention to the correct execution of the technique. | Please contact the corresponding author to request access to these images. |
| **L4. Knee flexion**  Stand behind a sturdy chair, resting your hands on the back of the chair or on the edge of the pool (water-based) to help balance. Stand straight, legs slightly apart, shift weight to one leg and, keeping balance, bend the working leg to bring the heel up towards the buttocks as high as you can or until your calf is parallel to the floor. To do repetitions established on one leg and then repeat with the same number on the other side.  ADAPTATIONS, VARIATIONS AND PROGRESSIONS:  Land-based:   a. Without equipment and progress to performing with ankle weights.   Water-based:   a. Execution speed (from slowest to fastest).   *SPECIAL CONSIDERATIONS:  - Pay special attention to the correct execution of the technique. | Please contact the corresponding author to request access to these images. |
| **5. Semi-Squat**  Stand with your feet at shoulder width, or slightly wider and the toes pointed slightly outward. Initiate the movement by sending the hips back as if you’re sitting back into an invisible chair. Bend knees to lower down with chest lifted in a controlled movement. Keep lower back neutral. Press through heels to stand back up to the starting position.  ADAPTATIONS, VARIATIONS AND PROGRESSIONS:   - As progression, go deep with more knee’s flexion. - As variation, sumo squat. Start by standing with your feet out wide and your toes pointing out.   Land-based:   a. If functional capacity is reduced, start with chair squat (sitting in a chair) or hold on to something solid, such as a chair or wall. Move on to performing without support when possible. b. Without equipment and progress to do it with dumbbells resting on hips.   Water-based:   a. Use one-hand support on the edge of the pool if necessary. b. Execution speed (from slowest to fastest).   *SPECIAL CONSIDERATIONS:  - Initiate the movement form the hip, not the knee  - Knees should not cross the toe | Please contact the corresponding author to request access to these images. |
| **L6. Adapted lateral squat**  Start with your feet wider than your hips and knees and toes pointing to the same direction, aligned. Shift your weight into your right heel, push your hips back, and bend that knee while leaving your left leg straight. Try to get your thigh parallel to the floor. Then, drive through your right foot to reverse the movement. Pause at the top to squeeze your glutes and stretch the front of your hips forward. Repeat on the other side.  ADAPTATIONS, VARIATIONS AND PROGRESSIONS:  Land-based:   a. If functional capacity is reduced, hold on to something solid, such as a chair or wall. Move on to performing without support when possible. b. Without equipment and progress to do it with dumbbells by holding it at your chest as you perform your reps.   Water-based:   a. Use one-hand support on the edge of the pool if necessary. b. Execution speed (from slowest to fastest). | Please contact the corresponding author to request access to these images. |
| **L7. Deadlifts**  Standing with your feet shoulder-width apart. Perform a hip flexion and lower the trunk under control with a light knee’s flexion. Come back to the initial position by driving your hips forwards, keeping a flat back.  ADAPTATIONS, VARIATIONS AND PROGRESSIONS:  Land-based:   a. If functional capacity is reduced, hold on to something solid, such as a chair or wall. Move on to performing without support when possible. b. Without equipment, with a dumbbell in each hand or with a resistance band holding the resistance band in both hands, loop the center around the base of your feet and stretching it by extending the hip.   Water-based:   a. Use one-hand support on the edge of the pool if necessary. b. With equipment that generates resistance when extending the hip, such as a board. | Please contact the corresponding author to request access to these images. |
| **L8. Split squat or lunge**  Start in a standing position with your feet hip-width apart. Step backward longer than a walking stride so one leg remains ahead of your torso and the other behind it. Your back foot should land at the ball of your foot with your heel lifted. Bend your knees to approximately 90 degrees as you lower yourself. Remember to keep your trunk upright and your hips level. Forcefully push off from the ball of the back foot to return to the starting position.  ADAPTATIONS, VARIATIONS AND PROGRESSIONS:  Variation: reverse lunge  Variation: forward lunge  Variation: Walking lunge  Land-based:   a. If functional capacity is reduced, hold on to something solid, such as a chair or wall. Move on to performing without support when possible. b. Without equipment and progress to do it with dumbbells adding weight.   Water-based:   a. Use one-hand support on the edge of the pool if necessary. b. Execution speed (from slowest to fastest).   *SPECIAL CONSIDERATIONS:  - Pay special attention to the correct execution of the technique. | Please contact the corresponding author to request access to these images. |
| **L9. Leg extension**  Land-based:  Lay down on and exercise mat with your feet flat on the floor and your knees pointing upwards bent at 90 degree angle. Lift one knee towards your chest and, holding the resistance band in both hands, loop the center around the base of your foot. Contract your glutes and extend your leg outwards at 45 degree angle until your knee is almost straight. Hold this extended position for 1 second before allowing your leg to be pulled back into its starting position. Do all reps on one side before switching.  Water-based:  Start in a standing position with your feet hip-width apart. Step on a pool noodle in the middle with one of the feet and drive working knee up to hip height. Extend the hip and knee sinking the pool noodle and making the effort during the extension. Return to the start and repeat right away. Do all reps on one side before switching. | Please contact the corresponding author to request access to these images. |
| **L10. Lying down bicycle exercise**  Land-based:  Lay down on, lift legs and bend knees so they are at a 90-degree angle, then begin rotating legs in a manner similar to riding a bicycle.  ADAPTATIONS, VARIATIONS AND PROGRESSIONS:   - Reduce the angle of the legs with respect to the ground, moving away from the vertical. - Use of ankle weights   Water-based:  Start from the body in supine flotation with the help of flotation equipment or clinging to the edge of the pool. Bend knees so they are at a 90-degree angle, then begin rotating legs in a manner similar to riding a bicycle.  ADAPTATIONS, VARIATIONS AND PROGRESSIONS:   - Execution speed (from slowest to fastest). | Please contact the corresponding author to request access to these images. |
| **L11. Standing calf raises**  Start in a standing position. Double heel raise using only body weight. Stand with equal weight on both feet. Raise both heels. Lower both heels simultaneously in a controlled manner.  ADAPTATIONS, VARIATIONS AND PROGRESSIONS:   - Unilateral hell raise   Land-based:   a. If functional capacity is severely reduced, it can be adapted to a sitting position or stand hold on to something solid, such as a chair or wall. b. Without equipment or with dumbbells in each hand to increase weight.   Water-based:   a. Use one-hand support on the edge of the pool if necessary. b. Execution speed (from slowest to fastest). | Please contact the corresponding author to request access to these images. |
| GENERAL CONSIDERATIONS AND RECOMMENDATIONS | |
| - Concentrate on proper form rather than rapid execution. Lift the weights with a smooth motion. - Select dumbbells of a weight you can lift to reach the RPE stablish without pain. Suggested starting without dumbbell or resistance bands (for land-based). - Exhale while lifting. - Pay special attention to the correct execution of the technique | |

Note. The images belong to a woman who participated in the program and a trainer of the program.
